# Improvement of pancreatic visualization in abdominal ultrasonography using milk tea ingestion with positional changes

**DOI:** 10.1101/2025.09.04.25334981

**Authors:** Ikuko Kawagishi, Hatsuka Ito, Yuri Ito, Yukimi Kojima, Shinpei Goto, Sesiru Takahashi, Kazunori Ohnishi

**Author notes:** Corresponding author: Kazunori Ohnishi, M.D., Department of Clinical Laboratory, Faculty of Medical Science, Shubun University, Nikko-cho 6, Ichinomiya, Aichi, 491-0938, Japan.

## Abstract

**Background:** The mortality of pancreatic cancer continues to rise, underscoring the importance of ultrasonography (US) for early detection in asymptomatic individuals. However, detection rates in screening settings remain unsatisfactory. While the liquid-filled stomach method has long been applied to improve pancreatic visualization, recent studies have suggested potential benefits of milk tea ingestion.

**Objective:** This study aimed to evaluate the effectiveness of gastric filling with milk tea combined with positional changes in enhancing pancreatic visualization on US.

**Methods:** Thirty participants (8 men, 22 women; mean age, 26.5 ± 14.5 years; median, 21; range, 18–75) were recruited, including healthy university students, family members, and staff. US examinations were performed using a Fukuda Denshi Sefius UF-890 AG by five trained students, following the standardized protocol of the Japan Society of Ultrasonics in Medicine. Before and after ingestion of 350 mL of milk tea, the long- and short-axis diameters of the pancreas, main pancreatic duct diameter, and skin-to-pancreas distance were measured. Gallbladder dimensions and bile duct diameter were assessed, and visibility of the pancreatic tail at the splenic hilum was recorded. Overall visualization was graded using a 5-point scale.

**Results:** Visualization scores improved significantly in all positions after milk tea ingestion (p < 0.05). Long-axis length increased significantly in the right lateral decubitus (p = 0.024) and left lateral decubitus (p = 0.012) positions. Gallbladder long-axis diameter decreased significantly after ingestion (p = 0.001). Pancreatic tail visibility was enhanced in the right lateral decubitus position both before and after ingestion. No significant correlations were observed between body mass index or skin-to-pancreas distance and pancreatic or gallbladder parameters.

**Conclusions:** Milk tea ingestion significantly improved pancreatic visualization on US, confirmed by both subjective grading and objective measurements, although the magnitude of improvement was modest. The effect appears to result from gastric filling serving as an acoustic window in specific positions. Given its palatability and ease of administration, milk tea may serve as a practical alternative to the water-drinking method, particularly in cases with suboptimal baseline visualization.

## Introduction

The number of deaths from pancreatic cancer has been increasing annually. According to cancer statistics from the National Cancer Registry of Japan in 2023, the incidence of pancreatic cancer was 36.5 per 100,000 population, and the mortality was 33.1 per 100,000. It ranks fourth among all cancers in men and third in women, underscoring its high prevalence. The 5-year survival rate remains only 8.5%, and long-term survival is rarely achieved in advanced stages ^1^. In contrast, a national survey on cancer screening conducted by the Japan Society of Ningen Dock (Human Health Screening Association) reported that pancreatic cancer was detected in only 0.004%–0.006% of asymptomatic individuals undergoing screening in 2012, which represents approximately 15% of the estimated incidence rate ^2^.

Thus, detection of pancreatic cancer through population-based screening remains insufficient. Early diagnosis using noninvasive ultrasonography (US) in asymptomatic individuals is therefore critically important. Among the various methods developed to improve pancreatic visualization, the liquid-filled stomach method has long been recognized ^3,4^. More recently, several studies from Japan have reported the potential usefulness of milk tea ingestion before examination ^5,6^.

In this study, we investigated the effectiveness of gastric filling with milk tea combined with positional changes in improving visualization of the pancreas on US, with quantitative assessment of different pancreatic regions.

## Subjects and Methods

### Subjects

A total of 30 participants were enrolled: 24 healthy university students from Shubun University, 4 family members, and 2 university staff members. The study population consisted of 8 men and 22 women, with a mean age of 26.5 ± 14.5 years (median, 21; range, 18–75). This study was approved by the Ethics Committee of the Faculty of Medical Science, Shubun University (Approval No. R07-001), and written informed consent was obtained from all participants.

## Methods

US examinations were performed using the Fukuda Denshi Sefius UF-890 AG ultrasound system. Five trained students from the Faculty of Medical Sciences conducted the measurements in accordance with the standardized procedures recommended by the Japan Society of Ultrasonics in Medicine ^7^.

All participants fasted prior to the examination. Measurements were taken before and after ingestion of 350 mL of milk tea (Kirin Gogo-no-Kocha Milk Tea®). For the pancreas, the long- and short-axis diameters were measured in the longitudinal view, and the horizontal diameter, vertical diameter, main pancreatic duct diameter, and pancreatic tail (at the splenic hilum) were assessed in the transverse view. The above values and the distance from the skin surface to the pancreas was measured in four positions: supine, semi-sitting (Fowler’s), left lateral decubitus, and right lateral decubitus.

For the gallbladder, the long-axis diameter, short-axis diameter, and bile duct diameter were measured from the right subcostal view. In addition, pancreatic visibility was graded on a five-point scale: 1 = almost not visible; 2 = only the body is visible; 3 = head or tail poorly visualized; 4 = generally good (pancreatic duct not clearly seen); 5 = good.

Statistical analyses were performed using EZR software (version R4.3.1). Paired t-tests or Wilcoxon signed-rank tests were applied for comparisons, and Pearson’s correlation coefficient was used to assess associations.

## Results

### 1. Pancreatic measurements before and after milk tea ingestion

Table 1 summarizes pancreatic measurements across positions and their statistical analyses. Overall, most measurements increased after milk tea ingestion. Significant increases were observed for the short-axis vertical diameter in the supine position, the long-axis short diameter in the sitting position, the long-axis length in the left lateral decubitus position, and the short-axis horizontal diameter in the right lateral decubitus position.

**Table 1.** Pancreatic measurements in different positions before and after milk tea ingestion and results of statistical analysis

| Position | Parameter | | Before milk tea<br>(mean ± SD, mm) | After Milk Tea<br>(mean ± SD, mm) | p-value* |
| --- | --- | --- | --- | --- | --- |
| Supine | Longitudinal view | Long axis | 72.0 ± 18.7 | 74.6 ± 15.3 | 0.338 |
| | | Short axis | 9.7 ± 2.8 | 10.8 ± 3.1 | 0.165 |
| | Transverse view | Vertical diameter | 18.2 ± 5.6 | 21.5 ± 5.4 | 0.005 |
| | | Horizontal diameter | 34.5 ± 9.6 | 34.8 ± 8.6 | 0.787 |
| Semi-Sitting | Longitudinal view | Long axis | 67.5 ± 14.9 | 69.3 ± 14.9 | 0.324 |
| | | Short axis | 9.3 ± 3.5 | 11.3 ± 4.3 | 0.018 |
| | Transverse view | Vertical diameter | 20.3 ± 6.0 | 20.5 ± 6.9 | 0.964 |
| | | Horizontal diameter | 33.8 ± 8.2 | 36.5 ± 7.4 | 0.100 |
| Left Lateral<br>Decubitus | Longitudinal view | Long axis | 53.2 ± 10.8 | 61.3 ± 14.3 | 0.012 |
| | | Short axis | 12.7 ± 3.8 | 12.5 ± 8.9 | 0.136 |
| | Transverse view | Vertical diameter | 22.2 ± 4.9 | 23.0 ± 5.3 | 0.501 |
| | | Horizontal diameter | 38.8 ± 10.5 | 37.6 ± 10.4 | 0.509 |
| Right Lateral<br>Decubitus | Longitudinal view | Long axis | 69.4 ± 19.4 | 77.8 ± 16.5 | 0.024 |
| | | Short axis | 12.5 ± 3.8 | 12.1 ± 3.6 | 0.663 |
| | Transverse view | Vertical diameter | 22.7 ± 6.3 | 23.3 ± 6.7 | 0.572 |
| | | Horizontal diameter | 34.1 ± 8.4 | 38.4 ± 8.0 | 0.029 |

\* Paired t-test or Wilcoxon signed-rank test

Because the long-axis length in the longitudinal view is considered the most reliable indicator of pancreatic visualization, the effect of milk tea ingestion was primarily evaluated using this measurement (Figure 1). Long-axis length increased in all positions following ingestion, with significant increases in the left lateral decubitus (p = 0.012) and right lateral decubitus (p = 0.024) positions. Before ingestion, the largest long-axis values were observed in the supine position; after ingestion, the right lateral decubitus position yielded the greatest measurements (Figure 2). The pancreatic tail was most clearly visualized in the right lateral decubitus position both before and after ingestion. Figure 3 shows a representative case in which pancreatic visualization improved after milk tea ingestion. In the right lateral decubitus position, the pancreatic long-axis length increased from 48.1 mm to 77.4 mm, and the visual grading score improved from 3 to 4.

**Figure 1.**
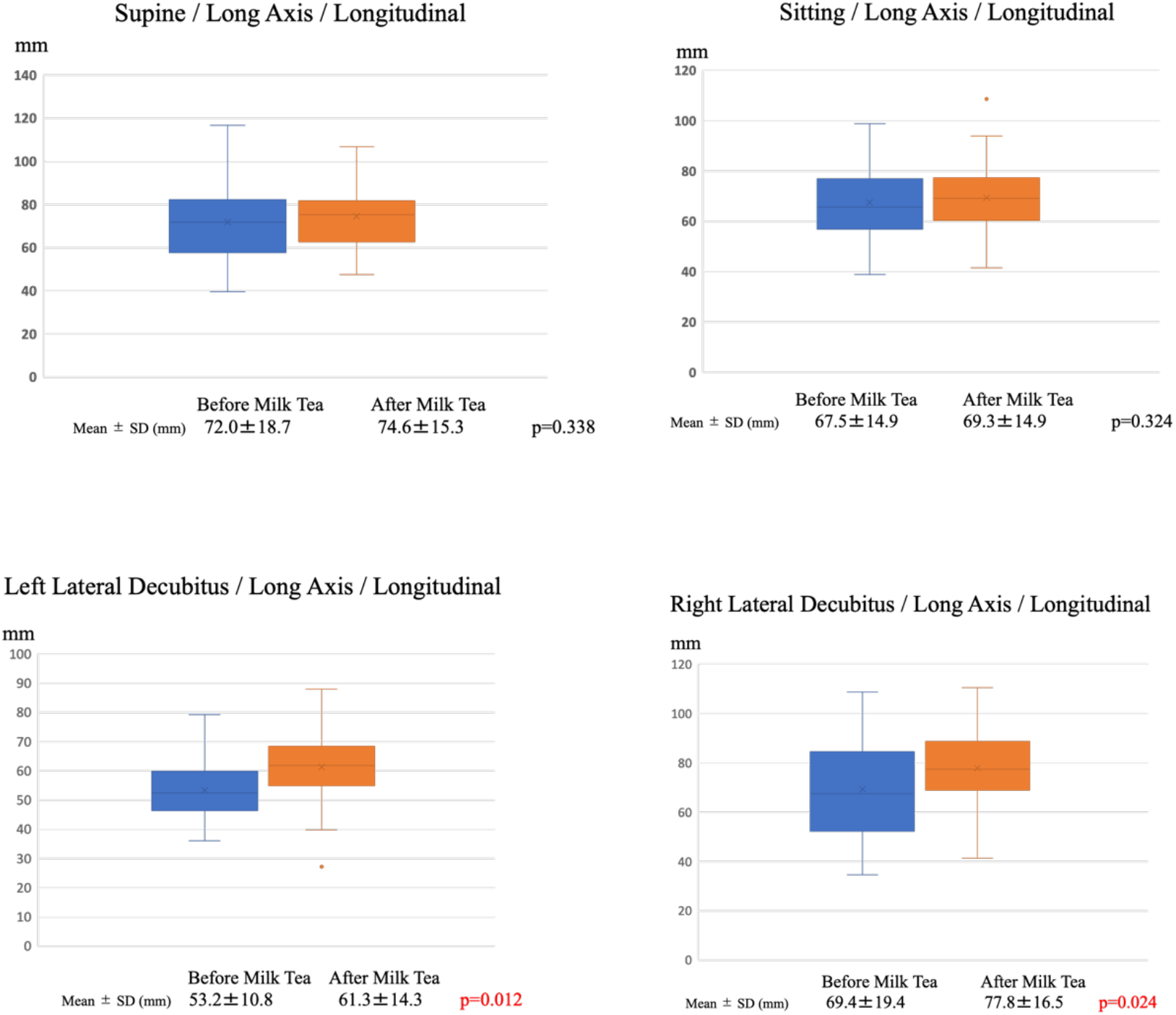
Changes in pancreatic long-axis length before and after milk tea ingestion across different positions

**Figure 2.**
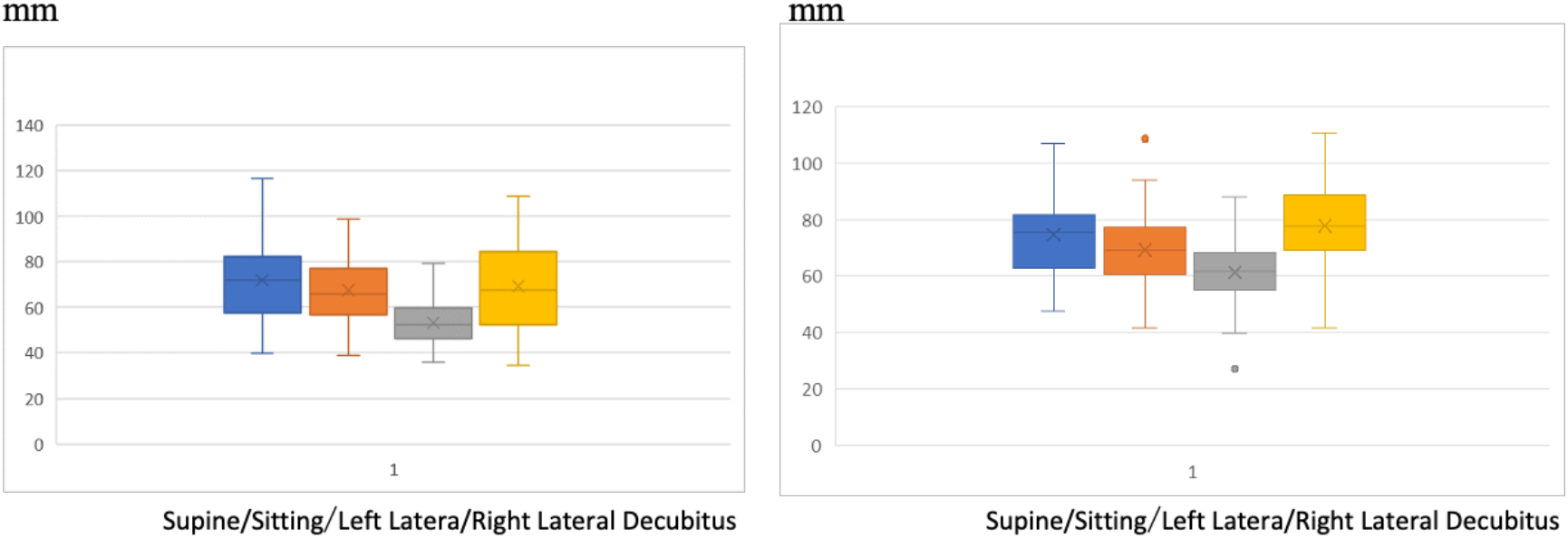
Comparison of pancreatic long-axis measurements in various positions before and after milk tea ingestion

**Figure 3.**
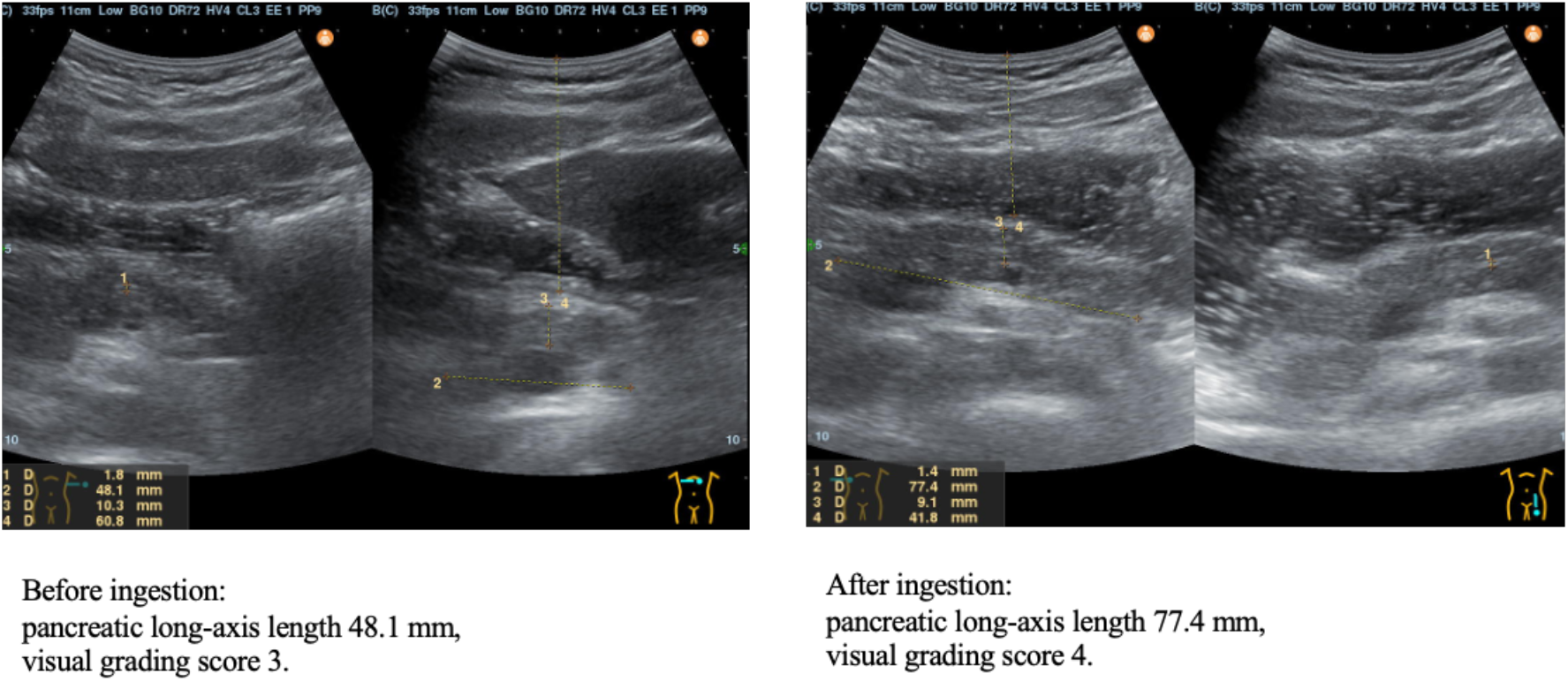
Pancreatic ultrasonography images in the right lateral decubitus position before and after milk tea ingestion.

### 2 Visual assessment of pancreatic visualization

On the 5-point grading scale, pancreatic visualization scores improved significantly in all positions following milk tea ingestion (Figure 4).

**Figure 4.**
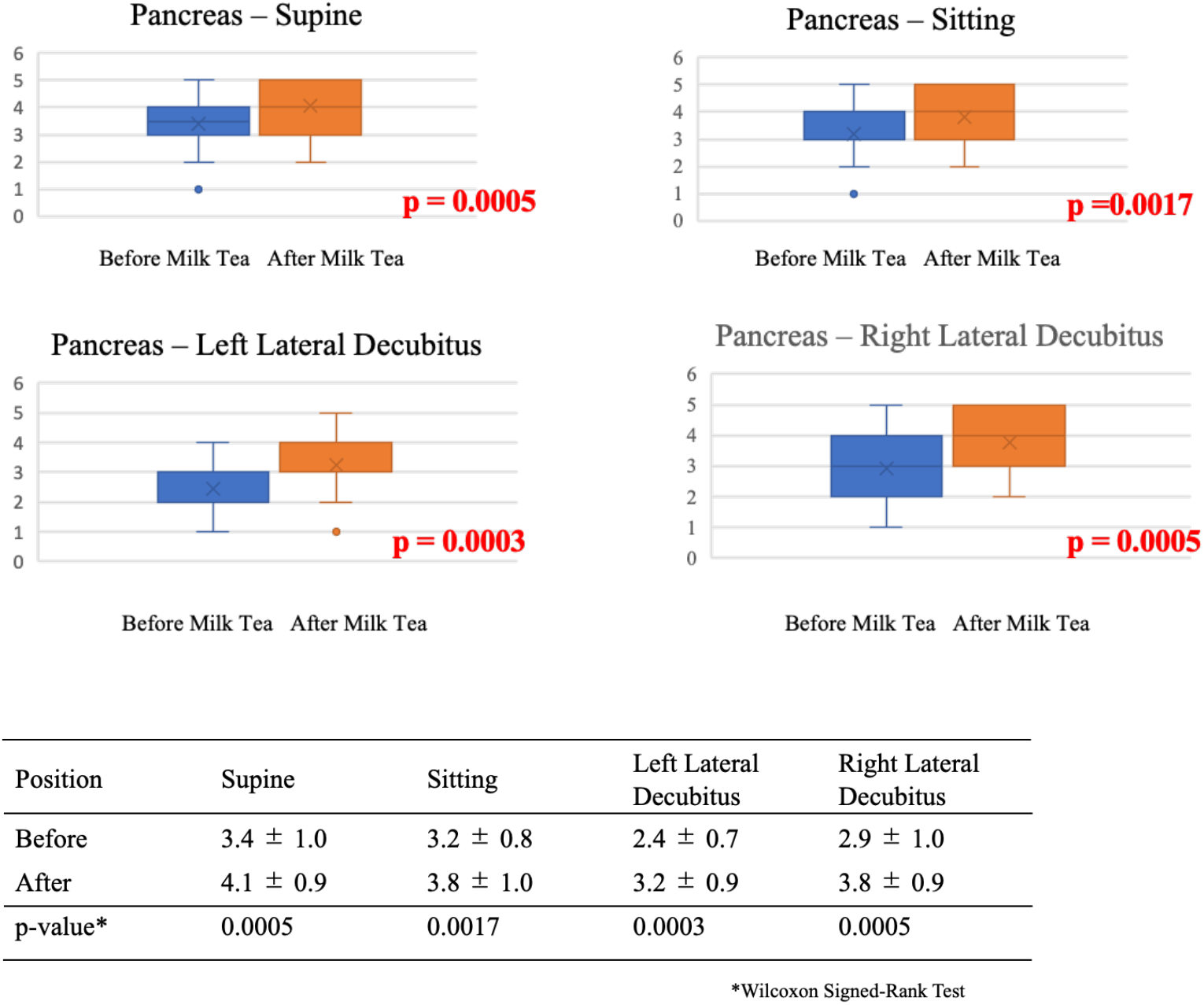
Visual grading scores of pancreatic visualization before and after milk tea ingestion

### 3. Relationship between pancreatic long-axis length and distance from the body surface

No significant correlations were observed between body mass index and any pancreatic or gallbladder measurement. Similarly, no correlation was found between the skin-to-pancreas distance and either pancreatic or gallbladder parameters (data not shown).

### 4. Gallbladder measurements before and after milk tea ingestion

The gallbladder long-axis diameter decreased significantly after milk tea ingestion (p = 0.001) (Figure 5). Gallbladder contraction was observed, and wall thickening was noted in three cases. All pancreatic and gallbladder measurements remained within normal limits: the maximum pancreatic short-axis diameter was <30 mm, no main pancreatic duct exceeded 3 mm, and no pancreatic masses or cysts were detected. For the gallbladder, no short-axis diameter exceeded 36 mm, and the bile duct diameter was <8 mm in all cases, with no evidence of dilatation or abnormal wall thickening.

**Figure 5.**
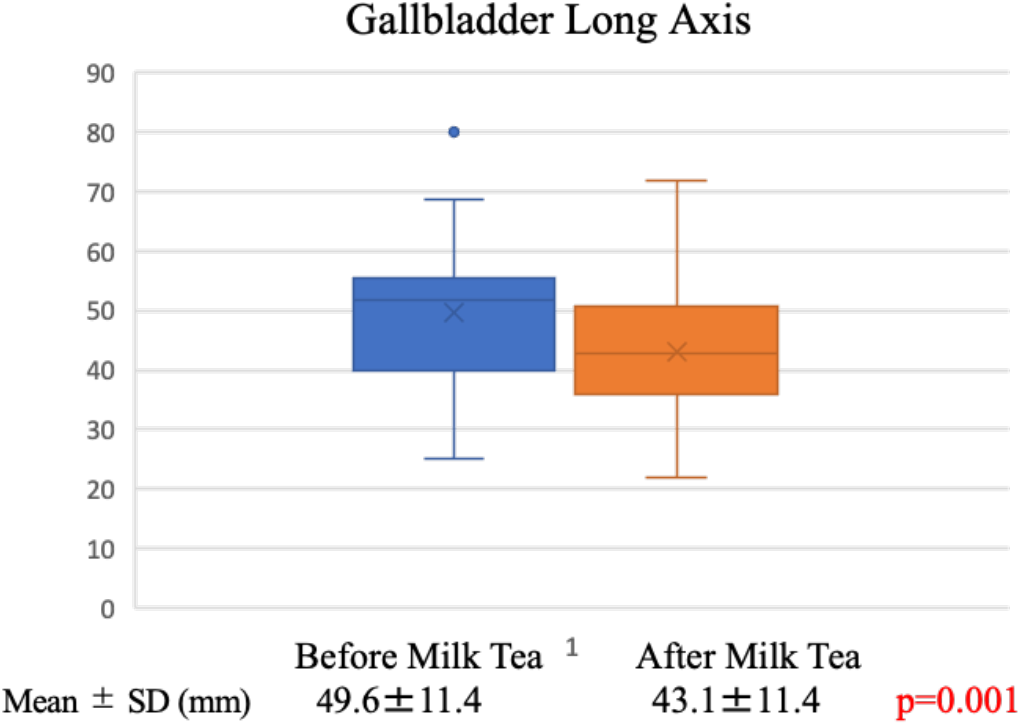
Gallbladder long-axis measurements before and after milk tea ingestion

## Discussion

This study demonstrated that milk tea ingestion improved pancreatic ultrasonography not only by visual assessment but also by quantitative measurements. Although the degree of improvement was modest, the findings support the use of milk tea as a simple and practical alternative to the conventional water-filled stomach method. The effect was most pronounced in the right lateral decubitus position, likely because gastric distension created a more effective acoustic window over the pancreas.

Anatomically, the pancreas lies in the retroperitoneal space, extending obliquely from the midline to the left posterior abdomen. Its deep and broad location makes complete visualization difficult. According to the 2023 National Cancer Registry of Japan, the overall 5-year survival rate for pancreatic cancer remains only 8.5%, with stage-specific survival rates of 42.1% for localized disease, 12.4% for regional disease, and 1.8% for distant metastases. These data underscore the importance of early detection during the localized stage ^1^.

Large-scale screening studies have identified several factors contributing to poor pancreatic visualization, including male sex, age ≥40 years, and BMI ≥25 ^7,8^. Moreover, the detection rate of pancreatic cancer during health checkups is reported to be only 0.004%–0.006%, highlighting the urgent need to improve diagnostic performance in screening ultrasonography ^8,9^. Ultrasound tends to underestimate pancreatic size compared with other imaging modalities. For example, one study reported a mean pancreatic body short-axis diameter of 12.7 ± 4.6 mm on ultrasound versus 14.5 ± 4.3 mm on MRI ^10^. This discrepancy further emphasizes the importance of techniques that enhance pancreatic visualization.

Several strategies have been proposed to improve pancreatic visualization in US. Positional changes can displace gastrointestinal gas and allow the liver to serve as an acoustic window; the semi-sitting position is advantageous for this reason. The pancreas, while retroperitoneal, is relatively mobile, making both lateral decubitus positions useful—particularly the right lateral decubitus for the body and tail and the left lateral decubitus for the head ^5,11^. Frequent repositioning also helps displace interfering gas.

Gastric gas commonly obscures the pancreatic body and tail in routine examinations. The liquid-filled stomach method, in which the stomach is distended with fluid to act as an acoustic window, has long been reported as effective ^3,4^. Studies from Japan have confirmed its utility using visual analog scales ^12^. More recently, milk tea has been evaluated as an alternative filling agent. Nakao et al. demonstrated significantly higher sensitivity for pancreatic cyst detection with pancreatic-focused US after milk tea ingestion (92.2%) compared with conventional abdominal US (70.2%), using MRI as the reference standard ^6^.

In our study, we quantitatively confirmed that milk tea ingestion improved pancreatic visualization. Although any clear liquid without fruit pulp, powdered tea, or bubbles can be suitable, palatability and ease of intake are critical. For instance, one study using 300 mL of tap water reported that 45% of participants could not finish the drink, suggesting that more acceptable beverages may be preferable ^12^. Milk tea, being familiar and well tolerated, may therefore improve compliance. For individuals with high risk for pancreatic cancer, additional diagnostic approaches are required, such as high-frequency probes and contrast-enhanced ultrasonography. When abnormalities such as hypoechoic masses, ductal dilatation, or strictures are detected during screening, further evaluation with endoscopic ultrasonography, CT, or MRI should be undertaken.

In conclusion, milk tea ingestion significantly improved pancreatic ultrasonography, as demonstrated by both visual grading and quantitative measurements, although the magnitude of improvement was modest. The benefit appears to derive from gastric distension serving as an acoustic window, particularly in the right lateral decubitus position. Given its palatability and convenience, milk tea may represent a practical alternative to the conventional water-filled stomach method, especially when baseline visualization is poor.

## Data Availability

All data produced in the present study are available upon reasonable request to the authors

